# Development of an effective immune response in adults with Down Syndrome after SARS-CoV-2 vaccination

**DOI:** 10.1101/2022.01.14.22269303

**Authors:** Laura Esparcia-Pinedo, Ayla Yarci-Carrión, Gloria Mateo-Jiménez, Noelia Ropero, Laura Gómez-Cabañas, Ángel Lancho-Sánchez, Enrique Martín-Gayo, Francisco Sanchez-Madrid, Fernando Moldenhauer, Ainhoa Gutiérrez-Cobos, Diego Real de Asúa, Arantzazu Alfranca

**Affiliations:** Immunology Department, Hospital Universitario de La Princesa, Instituto de Investigación Sanitaria Princesa, Madrid, Spain; Microbiology Department, Hospital Universitario de La Princesa, Instituto de Investigación Sanitaria Princesa, Madrid, Spain; Fundación de Investigación Biomédica del Hospital Universitario de La Princesa, Instituto de Investigación Sanitaria Princesa, Madrid, Spain; Biobanco, Fundación de Investigación del Hospital Universitario de La Princesa, Instituto de Investigación Sanitaria Princesa, Madrid, Spain; Department of Medicine, School of Medicine, Universidad Autónoma de Madrid, Madrid, Spain; Internal Medicine Department, Hospital Universitario de La Princesa, Instituto de Investigación Sanitaria Princesa, Madrid, Spain

## Abstract

Immune dysregulation in individuals with Down syndrome (DS) leads to an increased risk for hospitalization and death due to COVID-19 and may impair the generation of protective immunity after vaccine administration. The cellular and humoral responses of 55 DS patients who received a complete SARS-CoV-2 vaccination regime at one to three (V1) and six (V2) months were characterised. SARS-CoV-2-reactive CD4+ and CD8+ T lymphocytes with a predominant Th1 phenotype were observed at V1, and increased at V2. Likewise, a sustained increase of SARS-CoV-2-specific circulating Tfh (cTfh) cells was observed one to three months after vaccine administration. Specific IgG antibodies against SARS-CoV-2 S protein were detected in 96% and 98% of subjects at V1 and V2, respectively, though IgG titers decreased significantly between both timepoints.

**SUMMARY:** The work shows the cellular and humoral responses to SARS-CoV-2 vaccination of individuals with Down syndrome (DS) after one to three (V1) and six (V2) months. An effective immune response after six months was observed in 98% of DS individuals.

## INTRODUCTION

The new coronavirus disease (COVID-19) due to SARS-CoV-2 is a global health emergency since March 2020. Although COVID-19 entails no or mild symptoms in most individuals, several at-risk patient subgroups develop a severe form of the disease, leading to acute respiratory distress syndrome (ARDS) (Brodin, 2021; Ruan et al., 2020). Individuals with Down syndrome (DS), the most frequent chromosomal abnormality worldwide, are particularly vulnerable to the disease, with a 4-fold increased risk for COVID-19-related hospitalization and a 3 to 10-fold increased risk for COVID-19-related mortality (Clift et al., 2021; Hüls et al., 2021; Wadman, 2020).

Although this increase can be partially attributed to a higher prevalence of comorbidities associated with a poorer COVID-19-related prognosis, such as obesity, congenital heart disease (CHD) or respiratory diseases (Perera et al., 2020; Real de Asua et al., 2021) the susceptibility of this population to SARS-CoV-2 can be further explained by the immune dysregulation observed in DS. These individuals display immune features which promote the development of autoinflammatory and autoimmune conditions, and lead to an increase in the incidence of infections (Huggard et al., 2020). DS patients show higher levels of IFN-stimulated genes, which explain their higher basal levels of IFN-α signaling and hypersensitivity to IFN stimulation (Chung et al., 2021). Indeed, four subunits of IFN receptors are encoded on HSA21 (IFNAR1, IFNAR2, IFNGR2, and IL10RB), and an increase in IFNAR1 and IFNAR2 expression in different cell types from DS donors has been described (Kong et al., 2020). Cells from DS patients also present variable degrees of increased IFN-I proximal signaling, such as higher levels of pSTAT1 (Malle and Bogunovic, 2021). Furthermore, the increased expression of TMPRSS2, which primes the viral S protein for entry into host cells and is also encoded in HSA21, may facilitate the infection of target cells by SARS-CoV-2, while NLRP3 downregulation and increased IL-10 production could be involved in the higher risk of bacterial infectious complications in these patients (De Toma and Dierssen, 2021; Espinosa, 2020). DS individuals also have high basal levels of proinflammatory cytokines like IL-2, IL-6, or TNF-α, which may enhance the “cytokine storm” that underlies ARDS in this condition (Espinosa, 2020). Although the overall effect of these alterations seems deleterious and provides some proof for the worse outcomes following SARS-CoV-2 infection, the net balance between all these interplaying factors is far from clear. Indeed, a decrease in IFNAR2 has been associated with worse COVID-19 outcome (Pairo-Castineira et al., 2021), hence this mechanism would theoretically contribute to a better evolution of COVID-19 infections in DS patients.

This immune dysregulation may impair the generation of protective immunity after vaccine administration in individuals with DS. In fact, though people with DS usually develop protective antibody responses after vaccination, overall antibody levels tend to be lower and decline faster than in individuals without DS (Joshi et al., 2011). Due to their excess mortality risk and these concerns over vaccine protection, most countries have included adults with DS, especially those over 40 years, in prioritized vaccination groups (Hüls et al., 2020). However, data on the immune response elicited by SARS-CoV-2 vaccines in the DS population are lacking, and the degree of protection achieved is unknown. A thorough characterization of the humoral and cellular immunity mechanisms underlying the response of individuals with DS to COVID-19 vaccines is needed to appropriately tailor future vaccination campaigns for this vulnerable population. Moreover, this knowledge may also shed light onto general immune mechanisms involved in the response to SARS-CoV-2, and in the development of post-vaccination responses.

We performed a characterization of cellular and humoral responses developed in DS patients at one to three (V1) and six (V2) months after two-dose SARS-CoV-2 vaccination. We provide evidence of the presence of SARS-CoV-2-reactive CD4+ and CD8+ T lymphocytes with a predominant Th1 phenotype in V1, which increased in V2. Likewise, a sustained increase of SARS-CoV-2-specific circulating Tfh (cTfh) cells was observed in these patients one to three months after vaccine administration. Specific IgG antibodies against SARS-CoV-2 S protein were detected in V1, which significantly decreased in V2.

## RESULTS AND DISCUSSION

### T cell response to SARS-CoV vaccination in DS donors

To assess immune response elicited by SARS-CoV-2 vaccine in DS, we recruited 55 donors whose demographic and clinical characteristics are shown in Table 1. These donors were inoculated with two doses of a SARS-CoV-2 vaccine following the recommended schedule (vaccine types, described in Table 1), and both T cell and antibody responses were analyzed before vaccine administration (V0), one to three months (V1) and six months (V2) after second vaccination. Fifty-one subjects (51/55, 93%) were enrolled after having received at least one vaccination dose. To avoid any delay in official vaccination strategies, we admitted the use in these cases of biobank-stored samples (from earlier than February 2020) as baseline (V0). Median time (interquartile range) from the last dose of the recommended vaccination schedule to the first blood extraction (V1) was 62 (42-70) days, and 159 (150-172) days to the second blood extraction (V2).

**TABLE 1.** Demographic and clinical characteristics of the study sample.

|  |  |
| --- | --- |
|  | n = 55 |
| Age (years) | 44 ± 10 |
| Sex (male) | 25 (44%) |
| Living situation |  |
| Family home | 31 (55%) |
| Supervised group home | 1 (2%) |
| Institutionalized | 11 (20%) |
| Degree of intellectual disability |  |
| Mild | 45 (80%) |
| Moderate | 9 (16%) |
| Severe | 2 (4%) |
| Comorbidities(&) |  |
| Skin conditions | 36 (64%) |
| Hypothyroidism | 29 (51%) |
| Gastrointestinal disorders | 21 (37,5%) |
| Obstructive sleep apnea | 19 (34%) |
| Congenital heart disease | 10 (18%) |
| Alzheimer's disease | 11 (20%) |
| Epilepsy | 2 (4%) |
| Vaccine type |  |
| BNT162b2 (Comirnaty®, Pfizer/BioNTech) | 48 (86%) |
| mRNA-1273 (Spikevax®, Moderna) | 7 (12,5%) |
| ChAdOx1 (Vaxzevria®, Oxford/Astra Zeneca) | 1 (1,5%) |
| Adverse reactions (#) |  |
| To first vaccine dose | 6 (11%) |
| To second vaccine dose | 6 (11%) |
| History of COVID-19 before vaccination | 14 (25%) |
| Vaccinated in the past 12 months(*) | 10 (18%) |

(&): Only comorbidities that have been linked to a worse prognosis of SARS-CoV-2 infection or to immune dysregulation are presented.
(#): Noted adverse reactions were all mild (involving either local pain, fever or malaise, lasting less than 24 h).
(\*): Other than vaccination against SARS-CoV-2 (i.e., influenza or pneumococcal vaccination)

We determined the presence of SARS-CoV-2-specific T cells in V0, V1 and V2. To assess the percentage of responsive T cells, we used an activation-induced marker (AIM) assay based on the detection of CD4+ OX40/CD137 double positive and CD8+ CD69/CD137 double positive cells, after 24h in the presence of peptide pools from SARS-CoV-2 (NCAP, S1, S2, RBD domain, VME1 and Mpro proteins for V0 and S1, S2, and RBD domain for V1 and V2). In all cases, we included a human actin-derived peptide pool as a negative control, and Staphylococcal enterotoxin B (SEB) as a positive control.

As shown in Fig.1, we found a predominant CD4+ response (73.21% patients with specific CD4+ *vs*.16.36% patients with specific CD8+ in V2), which is in line with previous data in non-DS individuals (Grifoni et al., 2020). Of note, CD4+ activation against S1 and S2 was observed in a single pre-pandemic sample from V0 (Fig. 1A). We did not detect SARS-CoV-2-specific CD8+ lymphocytes in this baseline visit (Fig. 1B). SARS-CoV-2 S-responsive CD4+ cells have also been previously detected in healthy non-DS donors, and their presence is explained by a possible cross-reaction with common cold coronaviruses or other pathogens which contain SARS-CoV-2 homologous sequences (Woldemeskel et al., 2020). Interestingly, unlike our DS donors, NCAP-specific CD4+T cells have been frequently identified in healthy non-DS donors (Mateus et al., 2020), a finding which has been attributed by some authors to former exposure to SARS-CoV-2, since no NCAP-responsive CD4+ cells have been observed in pre-pandemic samples (Sekine et al., 2020). Because most V0 samples in our study were collected before the beginning of the pandemic, previous asymptomatic contact with SARS-CoV-2 can be reliably ruled out. In addition, it is conceivable that T cells recognizing NCAP might display reduced functional avidity, which would impair their detection in our activation-dependent assay. This possibility could be further supported by a feasible defect in *in vitro* T cell activation in DS patients, however lymphocytes from these individuals have been shown to respond *in vitro* to SEB, CMV and VZV to a similar extent as non-DS donors (Schoch et al., 2017).

**Figure 1.**
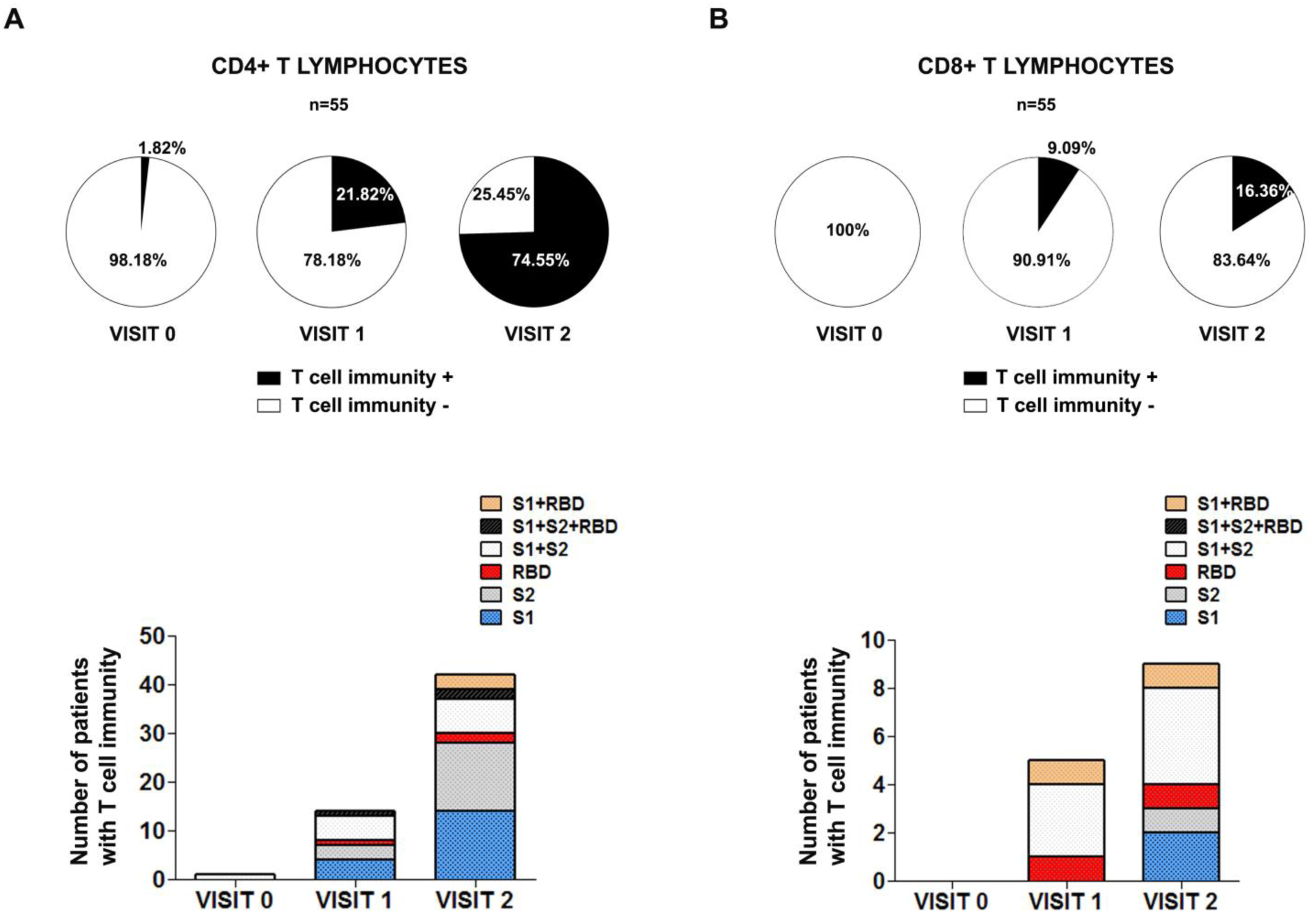
T cell response in Down Syndrome patients after SARS-CoV-2 vaccination. SARS-CoV-2-specific CD4+ (**A**.) and CD8+ (**B**.) lymphocytes in DS patients in Visit 0, Visit 1 and Visit 2. *Upper panels*, pie charts indicate the percentage of patients with (black) or without (white) T cell immunity against SARS-CoV-2 in each visit. *Lower panels*, number of patients with T cell response against specific peptide pools in each visit.

After vaccination, the percentage of individuals with SARS-CoV-2-responsive T lymphocytes raised from 23.8% in V1 to 73.21% in V2 for CD4+, and from 8.93% in V1 to 16.36% in V2 for CD8+. We could detect vaccine-elicited specific CD4+ and CD8+ directed against S1, S2 and RBD, with a similar pattern in V1 and V2 (Fig. 1A, B). T cell responses seem to be delayed in comparison to non-DS donors, which usually reach a higher percentage of individuals with specific T cell immunity within 4-7 weeks post SARS-CoV-2 vaccine administration (Esparcia-Pinedo L et al., 2022).

SARS-CoV-2 mRNA vaccines are known to promote a Th1 reaction in the general population (Sadarangani et al., 2021). Because individuals with DS usually present a higher Th1/Th2 ratio than euploid donors, a finding related to an increase in baseline IFNγ levels in this population (Franciotta et al., 2006), we sought to determine whether vaccination also resulted in Th1 predominant response in these patients. We evaluated T helper populations in SARS-CoV-2-specific CD4+ lymphocytes and found comparable percentages of Th1 and Th2 in V1. However, a significant increase in Th1 but not Th2 could be observed in V2 (p<0.01). As a specificity control, we determined Th1 and Th2 distribution in SEB-stimulated CD4+ cells, and found no significant differences between both subsets in either V1 and V2 (Fig. 2A). A similar pattern was detected regardless of the peptide pool (S1 or S2) analyzed (Fig. 2B). In addition, we did not find gender differences in Th1 and Th2 behavior, except for a significantly higher Th2 response in V1 in male DS donors (Suppl. Fig. 2A). Of note, although Th1 increase in V2 could be observed in all DS individuals, it was higher in patients ≤ 40 years of age (Suppl. Fig. 2B).

**Figure 2.**
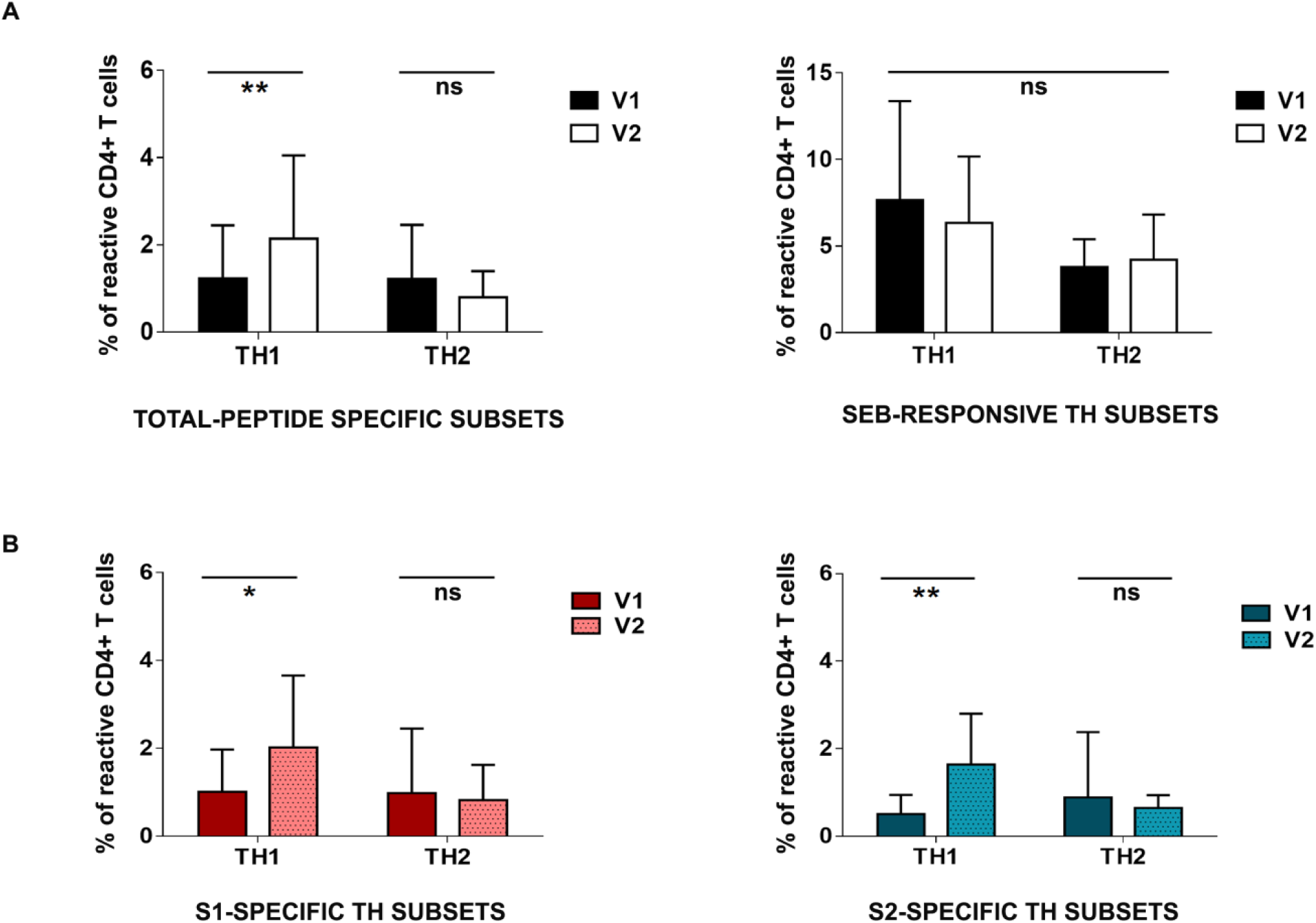
T helper subsets in Down Syndrome patients after SARS-CoV-2 vaccination. **A**., *Left*, percentage of SARS-CoV-2-specific Th1 and Th2 CD4+ lymphocytes in visits 1 (V1) and 2 (V2); *right*, percentage of SEB-activated Th1 and Th2 CD4+ cells in visits 1 and 2. **B**., Graphics show the percentage of S1-specific (*left*) and S2-specific (*right*) Th1 and Th2 CD4+ subsets in visits 1 and 2. Mean+SD is shown. *p<0.05; **p<0.01; ns, non-significant.

T follicular helper cells (Tfh) constitute a specialized subset of CD4+ T cells that collaborates in the activation and proliferation of B cells and the generation of high-affinity antibodies. In DS, normal counts of Tfh with a reduced B helper capacity have been reported. However other authors show increased percentages of IFNγ-secreting Tfh (Tfh1), which are thought to promote autoimmune responses in these patients (Ottaviano et al., 2020). In vaccinated DS donors, we found a SARS-CoV-2-specific cTfh population with sustained levels between V1 and V2 (Fig. 3A). PD-1hi cTfh are thought to represent a subset of recently activated cTfh cells (Dan et al., 2021). Accordingly, we detected SARS-CoV-2-specific CD4+CXCR5+PD-1hi cells in V1 in DS donors, whose levels were slightly reduced in V2 (Fig. 3A).

**Figure 3.**
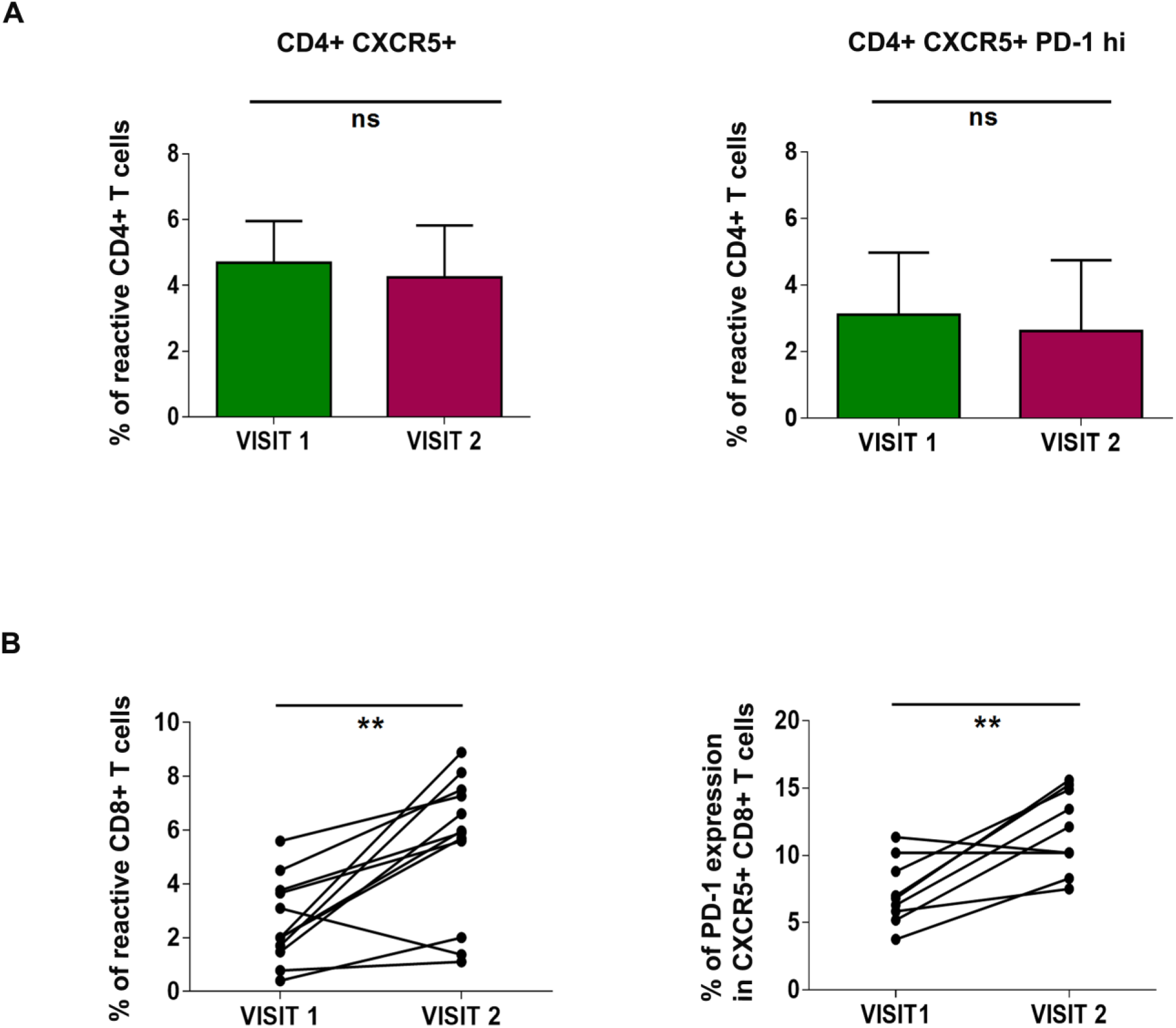
Circulating Tfh in Down Syndrome patients after SARS-CoV-2 vaccination. **A**., Graphics show the percentage of circulating CD4+CXCR5+ (*left*) and CD4+CXCR5+PD-1hi (*right*) cells in visits 1 and 2. Mean+SD is shown. **B**., Graphics show the percentage of SARS-CoV-2-specific CD8+CXCR5+ (*left*) and PD-1hi expression within CD8+CXCR5+ population (*right*) in visits 1 and 2 (n=11). *p<0.05; **p<0.01; ns, non-significant.

A population of CD8+CXCR5+ cells has been described which includes several functional subsets. Among them, a CD8+CXCR5+PD-1hi subset with Tfh-like properties displays cytotoxic characteristics in germinal centers, and may support antibody production in chronic infections and autoimmune conditions (Valentine and Hoyer, 2019). This population has been recently reported to increase in acute immunization settings and collaborate in specific antibody response (Tyllis et al., 2021). We therefore investigated the presence of SARS-CoV-2 specific CD8+CXCR5+PD-1hi cells in vaccinated DS patients and found that these cells were already detectable in V1, and their levels significantly increased in V2 (p<0.01) (Fig. 3B), suggesting that this cell subset could also cooperate in antibody generation in DS individuals after vaccine administration, although the actual functional significance of this population remains to be determined. In sum, our results indicate that SARS-CoV-2 vaccination efficiently induces the differentiation of specific populations that collaborate in an effective antibody response.

### Humoral response to SARS-CoV-2 vaccination in DS individuals

DS patients exhibit alterations in B lymphocyte maturation and function that may account for an impaired humoral response to vaccination. Transitional and mature naïve B cells are usually reduced by 50%, while numbers of switched memory B cells reach 10% of those found in non-DS counterparts (Carsetti et al., 2015). Similarly, a reduced frequency of switched memory B cells specific to vaccine antigens has been described in DS (Valentini et al., 2015). However, the degree of humoral response in DS individuals is highly variable among different vaccines. In this regard, influenza A/H1N1, pneumococcal capsular polysaccharide, or hepatitis B virus vaccination only induce a partial antibody response (Costa-Carvalho et al., 2006; Eijsvoogel et al., 2017), while conjugated pneumococcal or Hepatitis A virus vaccines have been reported to elicit adequate antibody titers. In our study, no patients were positive for anti-SARS-CoV-2 IgG before vaccination. We did find detectable titers of SARS-CoV-2 S protein specific IgG in 96.4% of patients (53/55) at V1. Two patients did not develop anti-S IgG antibodies at V1, one of which showed detectable titers of IgG at V2. The percentage of positive donors was similar at V2, when detectable titers of S protein-specific IgG were observed in 98.1% of patients (52/53, two patients with no data on humoral response at this timepoint; Fig.4A and Supplementary Table 1).

A significant decrease in IgG titers was found in 92.6% of patients (50/54) between both timepoints (Fig. 4B and Supplementary Table 1). Average specific IgG titers declined from 1520.84 BAU/mL in V1 to 780.88 BAU/mL in V2. This decrease was observed in the whole sample, though it was only statistically significant in the subgroup of individuals older than 40 years (Fig. 4C). Our results show an uncoupled humoral and cellular response in these patients, with antibody titers peaking already at V1, whereas T cell activation peaks at V2 (Fig.4). A dissociation between both arms of adaptive immunity has also been described in euploid individuals infected with SARS-CoV-2, and the development of an overt antibody response is not always accompanied by detectable T cell specific activation (Grifoni et al., 2020).

**Figure 4.**
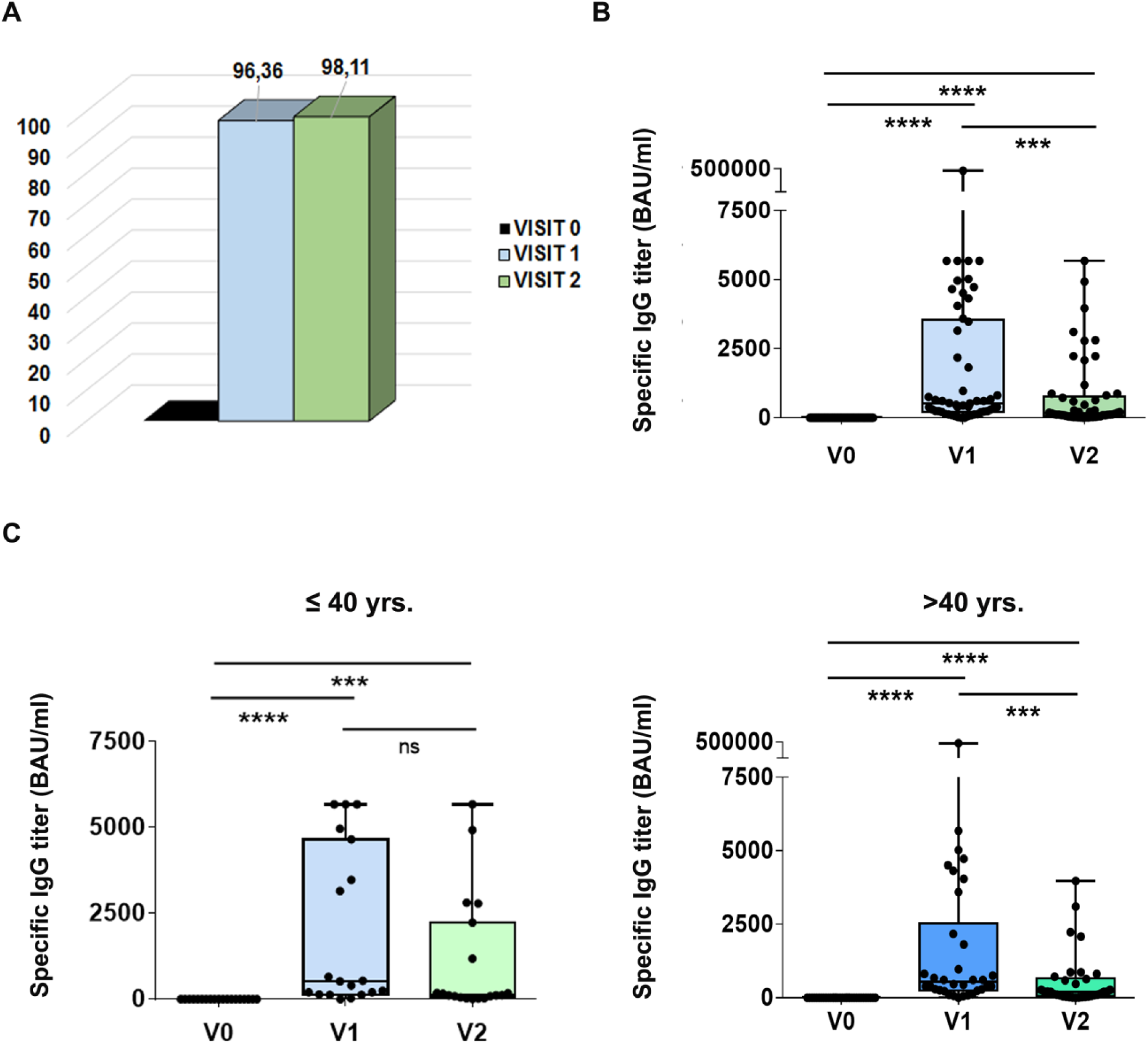
Humoral response in Down Syndrome patients after SARS-CoV-2 vaccination. **A**., Percentage of DS donors with detectable anti-SARS-CoV-2 S IgG in visits 0, 1 and 2. **B**., Specific anti-SARS-CoV-2 S IgG titers in DS patients in visits 0 (V0), 1 (V1), and 2 (V2). **C**., Specific anti-SARS-CoV-2 S IgG titers in visits 0 (V0), 1 (V1), and 2 (V2) in DS patients ≤ 40 years (*left*) or> 40 years (*right*). ****p<0.0001; ***p<0.001; ns, non-significant.

Our findings show that an immune adaptive response is achieved in DS individuals three months after SARS-CoV-2 vaccination lasting, at least, up to six months after vaccination. The degree of protection conveyed by these vaccination-elicited immunological changes should nonetheless be carefully interpreted. Recent reports still show an increased hazard ratio for COVID-19 hospital admission (2.55-fold increase) and mortality rate (12.7 fold increase) in DS patients, despite vaccination with one or two doses of either ChAdOx1 nCoV-19 or BNT162b2 (Hippisley-Cox et al., 2021). It is likely that other mechanisms beyond viral infection, and probably related to the intrinsic proinflammatory status present in patients with DS, operate to lead to a worse outcome even in vaccinated individuals. These indirect findings support the generalization of current recommendations to promote booster doses in the DS population. Moreover, because we observed a more robust response to vaccination in individuals with DS younger than 40 years, we believe our results lend further support to the recommendation of prioritizing the administration of an additional vaccine booster dose preferably in adults with DS over 40 years.

Yet unexplored but interesting issues are whether and how long vaccine-elicited responses persist over time in these patients. We are currently collecting samples to assess the evolution of humoral end cellular immunity up to twelve months after vaccination. Meanwhile, DS individuals over 40 years are already being administered a third dose of mRNA SARS-CoV-2 vaccine in Spain at the time of this publication. Analyzing the impact of this additional booster dose in their immune response will also be of great interest.

## MATERIALS AND METHODS

### Study population

This descriptive study was conducted with serological samples obtained from SD donors at La Princesa University Hospital, a tertiary level hospital in Madrid. Research subjects were consecutively recruited from those attending the Adult Down Syndrome outpatient unit, who consented to vaccination against SARS-CoV-2 during the 2021 COVID-19 vaccination season. The recruitment period ran from February 2021 to June 2021, and subject follow-up is ongoing at the time of this interim analysis.

Adults with DS over 18 years were considered eligible for participation in the study. DS diagnosis was established with a karyotype or compatible typical phenotype. Because the selection of a specific vaccine type (mRNA, adenoviral, etc) or regime depended on local availability and on the directives issued by the Spanish Ministry of Health -which have varied over time and between regions-, we determined that subject selection should not be limited to any single vaccine type. Inclusion in this study did not influence vaccination eligibility or regime, nor did it play a part in the usual clinical care received by adults with DS.

Patients were not eligible to participate in the study if they had a history of prior anaphylactic reaction to other vaccines/vaccine components or had were limited in their mobility and had strong difficulties to complete follow-up (i.e., many individuals living in residential settings).

### Detection of specific anti-SARS-CoV-2 IgG

All the samples were frozen at −20ºC after extraction. This is a standard of care that allows future serological assessments in case of need. Antibody titers were obtained by *SARS CoV-2 IgG II QUANT Alinity (Abbott*^*®*^). This assay is an automated, two-step immunoassay for the quantitative determination of IgG antibodies to SARS-CoV-2 against the spike receptor-binding domain (RBD) of SARS-CoV-2 in human serum using chemiluminescent microparticle immunoassay (CMIA).

SARS-CoV-2 IgG II Quant assay has demonstrated the ability to detect the spike RBD-based vaccine response in longitudinal samples from individuals both with and without prior COVID-19 infection (Ebinger et al., 2021; Narasimhan et al., 2021; Prendecki et al., 2021).

The interpretation of the results has been done following the criteria of the manufacturer, considering a negative result when a result <7,1 BAU/ mL was obtained. A positive result has been considered when a quantifiable result of ≥7.2 BAU/mL was obtained. The system has an analytical measurement range of 2,98-5680 BAU/mL. When there is > 5870 BAU/mL in the serum, the system reports it as> 5870 BAU/mL. The company reports a sensitivity of 99.3% and a specificity of 99.5% for this assay. We have expressed our results in BAU/mL (Binding antibody unit/mL). In line with the WHO International Standard study, the mathematical relationship of Abbott AU/mL unit to WHO BAU/mL unit would follow the equation: BAU/mL = 0.142 × AU/mL (STANDARDIZATION).

### PBMC Isolation and Culture

Peripheral blood mononuclear cells (PBMCs) were isolated using Ficoll-Paque (Pan-Biotech), following manufacturer’s instructions, and cryopreserved in liquid N2 in fetal bovine serum (HyClone TM) containing 10% DMSO (Inilab) and 1% penicillin/streptomycin (GIBCO) until use. For flow cytometry assays, 2×10^5^ cells per well were cultured for 24h at 37ºC and 5%CO2 atmosphere, in the presence of different SARS-COV-2 peptide pools (PepMixTM; JPT Peptide Technologies). Pools included peptides from S1, S2 and RBD from spike (S) protein, VME1 (membrane protein), NCAP (nucleoprotein) and Mpro (Cys-like protease, nsp5) (1μg/ml) in V0, and S1, S2 and RBD for V1/V2. A positive control with staphylococcal enterotoxin B (SEB; Sigma Aldrich) (1μg/ml), and a negative control with human β-actin peptide pool (PepMixTM; JPT Peptide Technologies) (1μg/ml) were included.

### Flow Cytometry

After stimulation, cells were incubated with anti-human CD3-PECy7/CD4-Pacific Orange/CD69-FITC/CXCR5-BV605/PD-1-BV786/CCR6-PerCP (BD Becton Dickinson)/CD137-BV711/OX40-PE/CXCR3-APCH7 (Biolegend) for 30 min. Subsequently, cells were washed and finally resuspended in 200μl PBS 1x. All the samples were acquired on a BD FACSCanto II flow cytometer (BD Becton Dickinson). The gating strategy for stimulated T cells is provided in Suppl Fig. 1: CD4+ T cells were identified as positive for CD3 and CD4 markers. Activated CD4+ cells were defined as double positive for OX40 and CD137. SARS-CoV-2-responsive Th1 and Th2 were defined as OX40+CD137+ cells within CXCR3+CCR6- and CXCR3-CCR6-subsets respectively. SARS-CoV-2-specific cTfh cells were defined as OX40+CD137+ double positive cells within CD4+CXCR5+ and CD4+CXCR5+PD-1hi populations. CD8+ T cells were identified as positive for CD3 and negative for CD4 staining, and activated CD8+ defined as CD69 and CD137 double positive cells. Flow cytometry data were analyzed using FlowJo software (BD Becton Dickinson).

Specific T cell response was determined by subtracting the percentage of either CD4+ OX40+CD137+ or CD8+ CD69+CD137+ in the presence of β-actin peptides from that obtained with SEB or SARS-CoV-2 peptides (specific percentage). A positive result was considered for samples with specific percentage equal or above the median two-fold standard deviation of all negative controls (0.82% for OX40/CD137 and 0.93% for CD69/CD137).

### Ethical considerations

This project was approved by the institutional IRB at La Princesa University Hospital (register number: 4386), meets international standards of data protection and is in line with general good practice guidelines and those established in the Declaration of Helsinki.

### Statistical analysis

Categorical data were described as frequencies (percentages) and quantitative variables, as mean (SD) or median (IQR). Quantitative variables were represented in bar graphs as mean ± standard deviation (SD), or in box-and-whiskers plots. For comparison between populations, analysis of variance was performed. To analyze statistically significant differences in variables following a non-normal distribution, Kruskal-Wallis test for unpaired samples and Friedman test for paired samples were used. The statistical significance threshold was set at p < 0·05. All data analyses were perfomed using Graph Pad Prism 8 Software (GraphPad Software, USA, www.graphpad.com), R (version 4.0.0), and SPSS.

### Online supplemental material

Supplemental material includes Supplemental Table 1 (detailed specific IgG titers in vaccinated DS patients), Supplemental Figure 1 (gating strategy for flow cytometry assays) and Supplemental Figure 2 (gender and age distribution of T helper subsets in vaccinated DS patients).

## Data Availability

All data produced in the present study are available upon reasonable request to the authors

## ACKNOWLEDGEMENTS

This work was supported by grants to A.A.: FIS PI19/01491 (Fondo de Investigación Sanitaria del Instituto de Salud Carlos III with co-funding from the Fondo Europeo de Desarrollo Regional FEDER) and Sociedad Cooperativa de Viviendas Buen Suceso, S.Coop.Mad. To A.A. and F.S-M.: CIBER Cardiovascular from the Instituto de Salud Carlos III (Fondo de Investigación Sanitaria del Instituto de Salud Carlos III with co-funding from the Fondo Europeo de Desarrollo Regional; FEDER). To F.S-M.: “Fondos Supera COVID19” (Banco de Santander and CRUE), “Ayuda Covid 2019” and “Inmunovacter” REACT-UE (Comunidad de Madrid). DRA was supported by the Fondo de Investigaciones Sanitarias (FIS grant PI19/00634, from the Ministerio de Economía y Competitividad (Instituto de Salud Carlos III) and co-funded by The European Regional Development Fund (ERDF) “A way to make Europe” and the Fondation Jérôme Lejeune (grant no. 2021a-2069). FM is supported by the Jérôme Lejeune Foundation (grant no. 2017b-1711). The Adult Down Syndrome Outpatient unit at Hospital Universitario de La Princesa is grateful to Licenciado don Jesús Coronado Hinojosa for his financial support of the research endeavors of our unit.

## COMPETING INTERESTS

The authors declare no competing financial interests.

## AUTHOR CONTRIBUTIONS

- Project coordination, original idea: AA, AG, DRA
- Subject recruitment and follow-up: FM, DRA
- Variable collection and database compilation: GM, AY, LE
- Initial sample processing, and sample collection preservation/storage: GM, AY, LE, NR
- Serological assays: AG, AY
- Cellular immunity activation assays: AA, LE, NR
- Lab coordination: AA, AG
- Statistical analysis and data mining: AG, AA, DRA
- Results evaluation, manuscript drafting and publication: All

## Notes

### Competing Interest Statement

The authors have declared no competing interest.

### Funding Statement

This work was supported by grants to A.A.: FIS PI19/01491 (Fondo de Investigacion Sanitaria del Instituto de Salud Carlos III with co-funding from the Fondo Europeo de Desarrollo Regional FEDER) and Sociedad Cooperativa de Viviendas Buen Suceso, S.Coop.Mad. To A.A. and F.S-M.: CIBER Cardiovascular from the Instituto de Salud Carlos III (Fondo de Investigacion Sanitaria del Instituto de Salud Carlos III with co-funding from the Fondo Europeo de Desarrollo Regional; FEDER). To F.S-M.: Fondos Supera COVID19 (Banco de Santander and CRUE), Ayuda Covid 2019 and Inmunovacter REACT-UE (Comunidad de Madrid). DRA was supported by the Fondo de Investigaciones Sanitarias (FIS grant PI19/00634, from the Ministerio de Economia y Competitividad (Instituto de Salud Carlos III) and co-funded by The European Regional Development Fund (ERDF) A way to make Europe and the Fondation Jerome Lejeune (grant no. 2021a-2069). FM is supported by the Jerome Lejeune Foundation (grant no. 2017b-1711). The Adult Down Syndrome Outpatient unit at Hospital Universitario de La Princesa is grateful to Licenciado don Jesus Coronado Hinojosa for his financial support of the research endeavors of our unit.

