## Supplemental material for "Development of an effective immune response in adults with Down Syndrome after SARS-CoV-2 vaccination"

#### Supplemental Table 1

| Specific IgG titer (BAU/mL) | | | | |
|---|---|---|---|---|
| | VISIT 1 | | VISIT 2 | |
| Patient | Titer | Interpretation | Titer | Interpretation |
| 1 | 247,61 | Ab detected | 184,07 | Ab detected |
| 2 | 615,40 | Ab detected | 93,61 | Ab detected |
| 3 | 206,07 | Ab detected | 118,70 | Ab detected |
| 4 | 3478,33 | Ab detected | 2231,87 | Ab detected |
| 5 | 29,61 | Ab detected | 24,79 | Ab detected |
| 6 | 1816,91 | Ab detected | 811,93 | Ab detected |
| 7 | 123,21 | Ab detected | | Missing |
| 8 | 63,53 | Ab detected | 204,18 | Ab detected |
| 9 | 70,33 | Ab detected | 10,17 | Ab detected |
| 10 | 4049,90 | Ab detected | 722,10 | Ab detected |
| 11 | 4512,85 | Ab detected | 3108,38 | Ab detected |
| 12 | **<7,1** | **Not detected** | 10,62 | Ab detected |
| 13 | >5680 | Ab detected | 1186,64 | Ab detected |
| 14 | 614,59 | Ab detected | 273,53 | Ab detected |
| 15 | 296,51 | Ab detected | 68,36 | Ab detected |
| 16 | 673,92 | Ab detected | 471,70 | Ab detected |
| 17 | 974,70 | Ab detected | 148,33 | Ab detected |
| 18 | 101,97 | Ab detected | 47,00 | Ab detected |
| 19 | 813,13 | Ab detected | 112,01 | Ab detected |
| 20 | 4320,44 | Ab detected | 595,63 | Ab detected |
| 21 | 384,92 | Ab detected | 57,41 | Ab detected |
| 22 | 403,14 | Ab detected | 52,16 | Ab detected |
| 23 | 2180,36 | Ab detected | 639,37 | Ab detected |
| 24 | 254,62 | Ab detected | 21,94 | Ab detected |
| 25 | 125,02 | Ab detected | 25,56 | Ab detected |
| 26 | 4729,92 | Ab detected | 872,19 | Ab detected |
| 27 | 4637.35 | Ab detected | 2235,09 | Ab detected |
| 28 | 365,54 | Ab detected | 35,00 | Ab detected |
| 29 | 470,79 | Ab detected | 38,16 | Ab detected |
| 30 | >5680 | Ab detected | >5680 | Ab detected |
| 31 | 3595,27 | Ab detected | 876,17 | Ab detected |
| 32 | 4657,00 | Ab detected | 4928,75 | Ab detected |
| 33 | 4965,24 | Ab detected | 2814,11 | Ab detected |
| 34 | 201,33 | Ab detected | 26,94 | Ab detected |

| | | | | |
|---|---|---|---|---|
| 35 | 3155,40 | Ab detected | 115,87 | Ab detected |
| 36 | 522,06 | Ab detected | 92,61 | Ab detected |
| 37 | >5680 | Ab detected | 2080,75 | Ab detected |
| 38 | 225,58 | Ab detected | 114,65 | Ab detected |
| 39 | 652,18 | Ab detected | 91,48 | Ab detected |
| 40 | 301,58 | Ab detected | 217,06 | Ab detected |
| 41 | 142,36 | Ab detected | 162,63 | Ab detected |
| 42 | 394,80 | Ab detected | 209,72 | Ab detected |
| 43 | 609,28 | Ab detected | 211,96 | Ab detected |
| 44 | 244,18 | Ab detected | 214,90 | Ab detected |
| 45 | 131,12 | Ab detected | 176,70 | Ab detected |
| 46 | 444,43 | Ab detected | 133,68 | Ab detected |
| 47 | >5680 | Ab detected | 2787,98 | Ab detected |
| 48 | 91,15 | Ab detected | 10,20 | Ab detected |
| 49 | 538,68 | Ab detected | 120,42 | Ab detected |
| 50 | 296,99 | Ab detected | | Missing |
| 51 | **<7,1** | **Not detected** | **<7,1** | **Not detected** |
| 52 | 5024,40 | Ab detected | 3971,39 | Ab detected |
| 53 | 757,64 | Ab detected | 270,71 | Ab detected |
| 54 | 392,62 | Ab detected | 100,78 | Ab detected |
| 55 | 125,02 | Ab detected | 25,56 | Ab detected |

**Supplemental Table 1. Titers of Specific IgG anti-SARS-CoV-2 S (BAU/ml) in DS patients at visits 1 and 2 after completing vaccination schedule.**

### SUPPLEMENTAL FIGURES

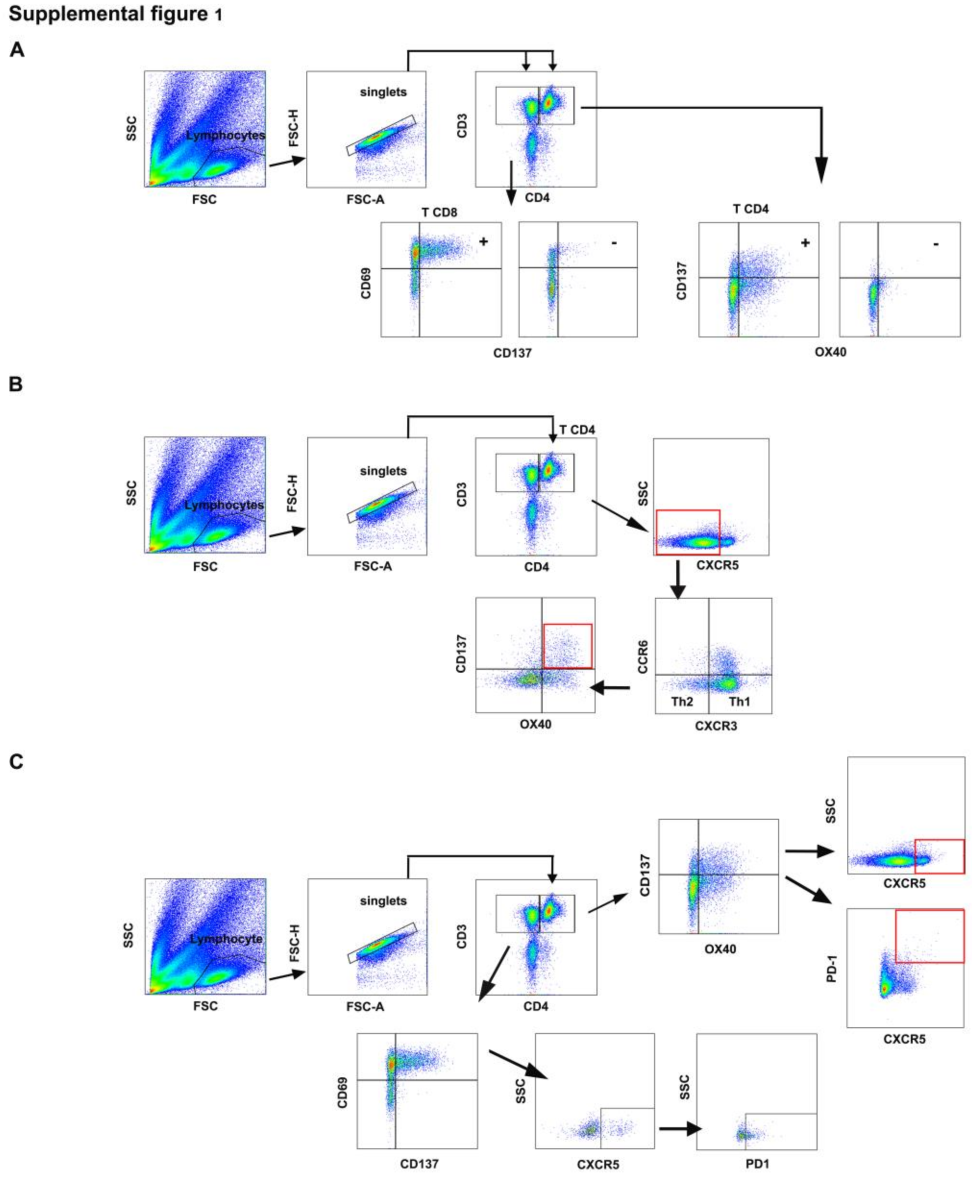


**Supplemental Figure 1. Gating strategy. A.,** Lymphocytes were gated in the FSC/SSC dot plot, followed by singlet gating in FSC-A/FSC-H dot plot. CD4+ T cells were identified in gated cells as CD3+CD4+. Activated CD4+ cells were defined as OX40+CD137+. CD8+ T cells were identified in gated cells as CD3+CD4-. Activated CD8+ cells were defined as CD69+CD137+. Dot plots show staining for both SEB-activated (+) and control β-actin-stimulated (-) lymphocytes. **B.,** SARS-CoV-2-specific Thelper subpopulations were defined as OX40+CD137+ cells within CD4+CXCR5-CXCR3+CCR6- (Th1) and CD4+CXCR5-CXCR3-CCR6- (Th2) subsets. **C.**, SARS-CoV-2-specific circulating Tfh cells were defined as CXCR5+ and CXCR5+PD-1hi cells within OX40+CD137+ CD4+ T lymphocytes, and SARS-CoV-2-specific CD8+CXCR5+ cells were defined as CXCR5+ and CXCR5+PD-1hi cells within CD69+CD137+ CD8+ T lymphocytes.

### Supplemental figure 2

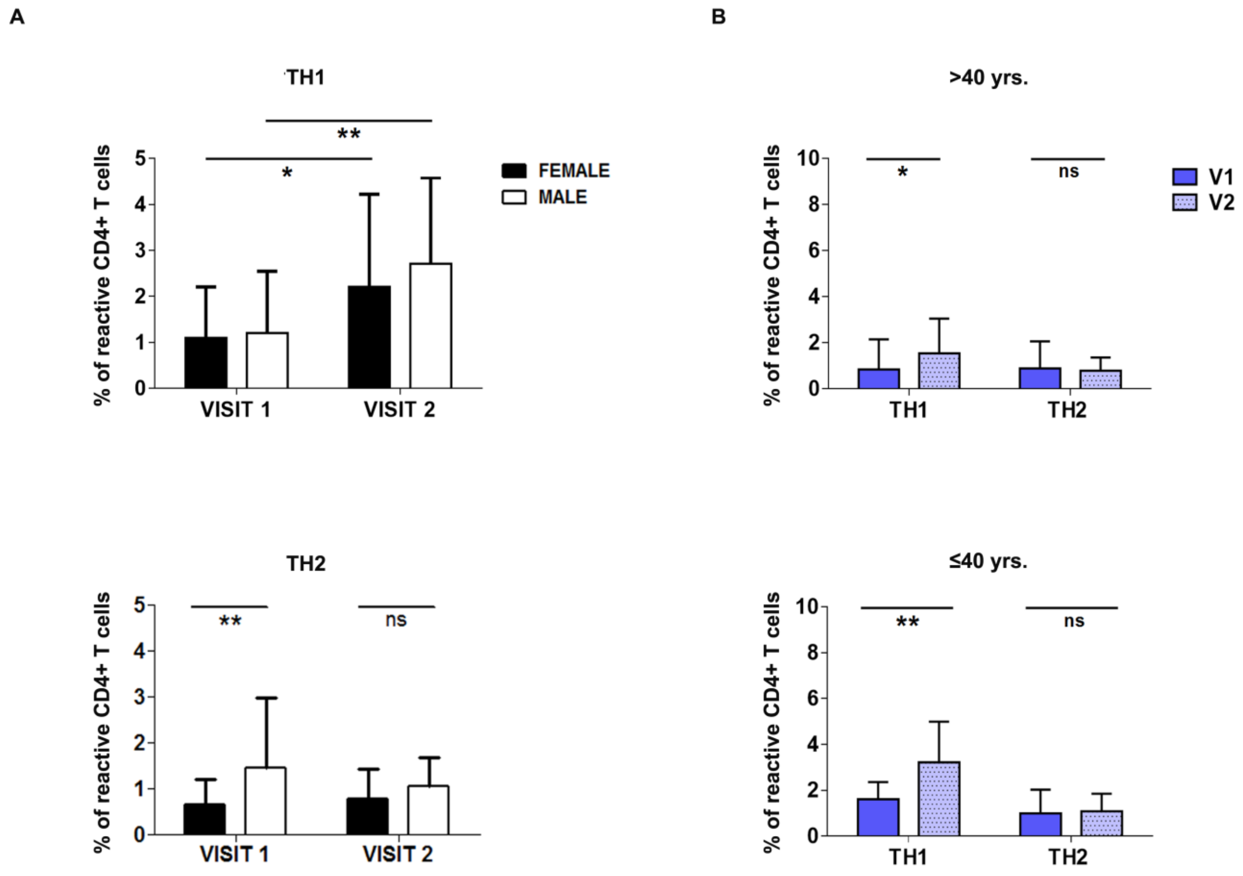


**Supplemental Figure 2. Gender and age distribution of T helper subsets in Down Syndrome patients after SARS-CoV-2 vaccination. A.**, Graphics show the percentage of SARS-CoV-2-reactive Th1 (*up*) and Th2 (*down*) from male (n=22) and female (n=29) patients in visits 1 and 2. **B.**, Percentages of SARS-CoV-2-reactive Th1 and Th2 in patients > 40 (n=30) (*up*) and ≤ 40 (n=11) (*down*) years in visits 1 (V1) and 2 (V2). Mean+SD is shown. *p<0.05; **p<0.01; ns, non-significant.
